# Effects of Smoking and Smoking Cessation on the Intestinal Microbiota

**DOI:** 10.1101/2020.07.11.20151480

**Authors:** Marcus G. Sublette, Tzu-Wen L. Cross, Claudia E. Korcarz, Kristin M. Hansen, Sofia M. Murga-Garrido, Stanley L. Hazen, Zeneng Wang, Madeline K. Oguss, Federico E. Rey, James H. Stein

## Abstract

**Introduction:** We evaluated associations of smoking heaviness markers and the effects of smoking cessation on the intestinal microbiota and cardiovascular disease risk factors in current smokers undertaking a quit attempt.

**Methods and Results:** Participants were current smokers enrolled in a randomized clinical trial of smoking cessation therapies with visits, risk factor measurements, and fecal collections at baseline, 2, and 12 weeks after starting a quit attempt. Genomic DNA was extracted from fecal samples followed by 16S rRNA gene sequencing and analysis using the QIIME2 software workflow. Relative abundances of bacterial taxa and alpha- and beta-diversity measures were compared.

Longitudinal changes in bacterial taxa abundances were compared using analysis of covariance (ANCOVA). The 36 smokers were (mean [standard deviation]) 51.5 (11.1) years old (42% male) and smoked 15.1 (6.4) cigarettes per day for 22.7 (11.9) pack-years. Their exhaled carbon monoxide (CO) levels were 17.6 (9.3) ppm. At baseline, relative abundances of the phylum *Actinobacteria* were correlated inversely with pack-years (rho=-0.44, p=0.008) and Cyanobacteria were correlated positively with CO levels (rho=0.39, p=0.021). After 12 weeks, abundances of the phyla*Bacteroidetes* increased (p_ANCOVA_=0.048) and *Firmicutes* decreased (p_ANCOVA_=0.036) among abstainers compared to continuing smokers. Increases in alpha-diversity were associated with lower heart rates (rho=-0.59, p=0.037), systolic blood pressures (rho=-0.58, p=0.043), and C-reactive protein levels (rho=-0.60, p=0.034).

**Conclusions:** Smoking cessation leads to minor changes in the intestinal microbiota. It is unclear if the proven health benefits of smoking cessation lead to salutary changes in the intestinal microbiota and if such changes affect cardiovascular disease risk.

**Implications:** In the largest prospective study of current smokers making a quit attempt to date, we showed that smoking cessation has minor effects on the composition of the gut microbiome. In successful abstainers, relative abundances of the phyla *Bacteroidetes* increased and *Firmicutes* decreased, a pattern of uncertain clinical significance. We did not observe significant changes in alpha- or beta-diversity with smoking cessation. It is unclear if the proven health benefits of smoking cessation lead to salutary changes in the intestinal microbiota and if such changes affect cardiovascular disease risk.

## INTRODUCTION

The gut microbiota appears to play a pathophysiologic role in atherosclerosis and cardiovascular disease, possibly through interactions with the immune system resulting in chronic systemic inflammation, by contributions to lipid metabolism, and/or through direct interactions of microbial-derived products such as trimethylamine N-oxide (TMAO) with vascular endothelium and platelets.[1,2] Cigarette smoke may modulate gut microbiota by upregulating oxidative stress-related enzymes in gut immune tissue, altering the gut mucin layer, expression of intestinal tight junction proteins, and local acid/base imbalance in the colon, through direct toxic effects from the myriad compounds in tobacco smoke, or through the spread of bacteria directly from cigarettes.[3,4] Although there is evidence that the intestinal microbiota of smokers differs from non-smokers,[5,6] it is not known whether smoking cessation changes the gut microbiota and how these changes may relate to pre-quit smoking heaviness, weight gain, and cardiovascular disease risk factors. A previous report described profound changes in the intestinal microbiota following smoking cessation, however that was a very small study and the observed microbiome changes were not integrated with vascular and inflammatory biomarkers.[3,5,6]

In this pilot study, we sought to identify plausible associations between smoking heaviness, metabolic markers, and the composition of the intestinal microbiota and their changes 12 weeks after a quit attempt. In contrast with the previous published work,[3,5,6] our results suggest that smoking cessation is associated with minor changes in the gut microbiome.

## METHODS

### Participants

The University of Wisconsin Health Sciences Institutional Review Board approved this study. This study was conducted in accordance with all University, State, and Federal research regulations. All participants provided free informed consent. Participants were recruited from the ongoing Quitting Using Intensive Treatment Study (QUITS) Trial (NCT03176784) and enrolled from February 19^th^ to August 16^th^, 2019. The main QUITS inclusion criteria were age ≥18 years old, smoking ≥10 cigarettes per day, and no use of pipe tobacco, cigars, snuff, e-cigarettes or chewing tobacco in the preceding 30 days. This sub-study also excluded individuals with factors known to affect the gut microbiota including: use of systemic antibiotics, corticosteroids, immunomodulators, or commercial probiotics in the last 6 months, major gastrointestinal surgery, and active inflammatory bowel disease or other gastrointestinal disorders.

### Design

All participants received varenicline (1 mg daily) and either placebo or nicotine patches (14 mg for 2 weeks pre-quit and then 10 weeks post-quit, then 7 mg patches for weeks 11 and 12 after the quit date). Smoking cessation was confirmed by 7-day self-report and verified by exhaled carbon monoxide (CO, Micro^+^™ Smokerlyzer^®^ CO monitor, Bedfont Scientific Ltd, Maidstone, Kent, England) levels. Participant measures were obtained at baseline (immediately prior to the quit attempt), 2 weeks post-quit attempt, and 12 weeks post-quit attempt.

### Processing and Analysis of Fecal Samples

Fresh stool samples were collected by participants and kept refrigerated (4-8B°C) until delivery to the microbiology lab where an aliquot of feces was immediately stored at −80°C until further processing. Fecal samples were kept refrigerated for an average of 3 hours and 43 minutes between collection to storage, ranging from 40 minutes to 8 hours and 5 minutes.

#### DNA extraction

Genomic DNA was extracted from fecal aliquots using a bead-beating protocol.[7] Briefly, feces was re-suspended in a solution containing 500 μl of extraction buffer [200 mM Tris (pH 8.0), 200 mM NaCL, 20 mM EDTA], 210 μl of 20% SDS, 500 μl phenol:chloroform:isoamyl alcohol (pH 7.9, 25:24:1, Invitrogen, Carlsbad, CA) and 500 μl of 0.1-mm diameter zirconia/silica beads. Samples were mechanically disrupted using a bead beater (BioSpec Products, Barlesville, OK; maximum setting for 3 minutes at room temperature), followed by centrifugation, recovery of the aqueous phase, and precipitation with sodium acetate and isopropanol.

QIAquick 96-well PCR Purification Kit (Qiagen, Germantown, MD) was used to remove contaminants. Isolated DNA was eluted in Tris-EDTA buffer and stored at −80 °C until further use.

#### Sequencing

PCR was performed using universal primers flanking the variable 4 (V4) region of the bacterial 16S rRNA gene.[16] PCR products were purified with the QIAquick 96-well PCR Purification Kit (Qiagen, Germantown, MD). Samples were quantified by Qubit Fluorometer (Invitrogen, Carlsbad, CA) and equimolar pooled. The pool was sequenced at the University of Wisconsin-Madison Biotechnology Center with the MiSeq 2×250 v2 kit (Illumina, San Diego, CA, USA). DNA extraction blanks, PCR blanks, and technical duplications for both extractions and PCRs were employed to ensure proper sample handling throughout library preparation process.

#### Analysis

Sequences were processed using QIIME 2 pipeline (2019.1).[8] Briefly, demultiplexed paired-end sequences were imported using Casava 1.8 format and denoised using DADA2[9] to obtain amplicon sequence variant (ASV) table. Singletons (ASV that were observed fewer than 2 times) and ASVs that are present in less than 10% of the samples were discarded. A naive Bayes taxonomy classifier was trained on the Greengenes[10] reference database (clustered at 99% similarity). This classifier was used to assign taxonomy to ASV.[11] ASV were then collapsed based on genus-level taxonomy. An even sampling depth (sequences per sample) of 4,135 sequences per sample was used for assessing alpha- and beta-diversity measures.

Pielou’s evenness index,[12] Faith’s phylogenetic diversity (PD), and Shannon diversity index were used to measure alpha diversity. Beta-diversity was calculated using Bray-Curtis, Jaccard, and weighted and unweighted UniFrac metrics.[13]

### Statistical Analysis

All analyses used XLStat (Addinsoft, France) in R (R Core Team, 2014). Distributions were inspected for zero values and non-linearity. Spearman’s rank correlations were used to compare relative abundances of microbial taxonomy, smoking heaviness markers, and other measures at baseline. ANCOVA was used to compare temporal changes in intestinal microbiota abundances and diversity measures by eventual cessation group (abstainers vs. continuing smokers) and each baseline measure. Given the exploratory nature of our study, we estimated statistical power using the genus taxonomic level with up to 13 different categories. With a sample of size of 30, we had 80% power to detect a rank correlation of 0.57 between the abundance metrics and outcome measures using a two-sided test for significance at a taxonomic family-wise 0.05 level, however we did not perform formal adjustments for multiple comparisons.

## RESULTS

### Participant Characteristics (Table 1)

The 36 smokers that attended the baseline visit were mean (standard deviation) 51.5 (11.1) years old, 58% female, and they smoked 15.1 (6.4) cigarettes per day for 22.7 (11.9) pack-years. Their exhaled CO levels were 17.6 (9.3) ppm. Greengenes classifier assigned usable raw reads to 11 phyla, 35 families, and 55 genera. The most abundant phyla were *Firmicutes* and *Bacteroidetes*.

**Table 1.** Participant Characteristics at Baseline (N=36)

| Variable | Mean | Standard Deviation | Range |
| --- | --- | --- | --- |
| Age (years) | 51.4 | 11.2 | 26 – 71 |
| Male (%) | 42.0 | - | - |
| Cigarettes/day | 15.1 | 6.4 | 5.1 - 35.0 |
| Pack-years | 22.7 | 12.0 | 2.5 - 52.0 |
| Exhale Carbon monoxide (ppm) | 17.6 | 9.3 | 5 – 41 |
| Body-mass index (kg/m<sup>2</sup>) | 29.6 | 8.4 | 20.7 - 67.9 |
| Heart rate (bpm) | 72.3 | 10.7 | 57 – 100 |
| Systolic blood pressure (mmHg) | 131.4 | 16.5 | 102 – 162 |
| Diastolic blood pressure (mmHg) | 77.6 | 9.0 | 66 – 95 |
| Non-high-density lipoprotein cholesterol | 160.3 | 52.6 | 69 – 274 |
| Glucose (mg/dL) | 92.4 | 9.7 | 70 – 109 |
| C-reactive protein (mg/L) | 4.6 | 7.5 | 0.1 - 39.5 |
| Trimethylamine N-oxide (microM) | 3.75 | 2.11 | 1.11 - 13.18 |
| White blood cell count (K/μL) | 8.4 | 2.2 | 4.6 - 13.9 |
| Carotid-femoral pulse wave velocity (m/s) | 7.46 | 1.84 | 4.80 – 12.95 |
| Brachial artery flow-mediated dilation (%) | 4.57 | 2.91 | 0.55 – 12.05 |
| Carotid plaque score (0-12) | 3.0 | 2.8 | 0 – 10 |
| **Microbiota Relative Abundances (%)** |  |  |  |
| *Actinobacteria* | 4.17 | 4.37 | 0.0 - 15.90 |
| *Cyanobacteria* | 0.04 | 0.13 | 0.0 - 0.72 |
| *Verrucomicrobia* | 0.70 | 2.55 | 0.0 - 14.41 |
| *Euryarchaeota* | 0.10 | 0.24 | 0.0 - 0.96 |
| *Bacteroidetes* | 23.40 | 10.5 | 8.10 - 49.30 |
| *Firmicutes* | 70.20 | 10.0 | 46.10 - 88.70 |
| *Fusobacteria* | 0.011 | 0.44 | 0.0 - 2.52 |
| *Proteobacteria* | 1.26 | 1.02 | 0.0 - 5.70 |
| **Alpha-Diversity Measures (units)** |  |  |  |
| Pielou's evenness | 0.77 | 0.06 | 0.58 - 0.84 |
| Faith PD | 8.32 | 1.68 | 4.78 - 12.11 |
| Shannon | 5.14 | 0.60 | 3.05 - 6.12 |

### Baseline Correlations between Microbial Diversity Scores, Bacterial Taxa Abundances and Smoking Measures

Pack-years of cigarettes smoked correlated inversely with relative abundance of *Actinobacteria* (rho=-0.44, p=0.008) and exhaled CO correlated positively with relative abundance of Cyanobacteria (rho=0.39, p=0.021). No significant associations between *Actinobacteria* or Cyanobacteria and smoking measures were identified, nor were taxa at the family or genus levels within these two phyla. The relative abundance of the *Actinobacteria* phylum correlated inversely with age (rho=-0.44, p=0.007) and carotid artery plaque score (rho=-0.33, p=0.049) and directly with glucose (rho=0.038, p=0.021). *Actinobacteria* and Cyanobacteria were not associated with TMAO, hsCRP, or any other cardiovascular disease risk measures. Exhaled CO was associated with the genus Dorea (rho=0.43 p=0.009), otherwise, none of the smoking heaviness measures were associated with other diversity index or bacterial abundances at the phyla, family, or genera levels.

### Effects of Smoking Cessation and Continued Smoking (Table 2, Supplementary Figure)

Of the 29 participants that returned for the week 2 visit, 17 had quit smoking. There were no significant between-groups differences in relative abundances of bacteria or diversity measures at this early time point. Of the 26 that returned for the week 12 visit, 14 successfully quit smoking. Those who quit had greater reductions in exhaled CO levels than continuing smokers (−14.0 [10.4] vs. +2.1 [7.6] ppm, p_ANCOVA_<0.001). Significant between group differences in changes were not observed for cardiovascular disease risk factors, hsCRP, TMAO, PWV, and brachial artery FMD. Significant between group differences were not observed for relative abundances of *Actinobacteria* (p_ANCOVA_=0.150) or *Cyanobacteria* (p_ANCOVA_=0.165). However, *Bacteroidetes* increased (p_ANCOVA_=0.048) and *Firmicutes* decreased (p_ANCOVA_=0.036) among successful abstainers compared to continuing smokers. No other significant changes between groups were identified for relative abundances of bacteria at the phylum level.

**Table 2.** Effect of Quit Attempt on Changes in Bacterial Abundance and Diversity Measures from Baseline to Week 12.

| **Change Variable** | **Beta** | **Standard Error** | **P-value** |
| --- | --- | --- | --- |
| **Microbiota Relative Abundances (%)** |  |  |  |
| *Actinobacteria* | 1.2 | 0.8 | 0.15 |
| *Cyanobacteria* | 0.0 | 0.0 | 0.17 |
| *Verrucomicrobia* | 0.0 | 0.1 | 0.92 |
| *Euryarchaeota* | 0.0 | 0.4 | 0.99 |
| *Bacteroidetes* | 7.4 | 3.5 | **0.048** |
| *Firmicutes* | -7.6 | 3.4 | **0.036** |
| *Fusobacteria* | -0.1 | 0.1 | 0.53 |
| *Proteobacteria* | 0.3 | 0.6 | 0.67 |
| **Alpha-Diversity Measures (units)** |  |  |  |
| Pielou's evenness | -0.01 | 0.02 | 0.61 |
| Faith PD | -0.10 | 0.85 | 0.91 |
| Shannon | -0.13 | 0.23 | 0.59 |

Within *Firmicutes*, an undefined family within the order *Clostridiales* had a significant difference in abundance between smoking status groups at week 12 (p_ANCOVA_=0.043). At the genus level, *Ruminococcus* (p_ANCOVA_=0.027) and an undefined genus within order *Clostridiales* (p_ANCOVA_=0.043) also had significant differences in abundances between smoking status groups. Although changes in alpha- and beta-diversity measures over time did not differ between successful abstainers and continuing smokers, increases in alpha-diversity 12 weeks following a smoking cessation attempt were associated with lower heart rates (rho=-0.59, p=0.037), systolic blood pressures (rho=-0.58, p=0.043), and C-reactive protein (rho=-0.60, p=0.034).

As expected, nicotine patch users were more successful at quitting with greater reductions in CO at week 12 visit (p=0.006), however there were no significant differences in the gut microbiota between participants that received a placebo or a nicotine patch.

## Discussion

In this pilot study of daily cigarette smokers, pack-years of smoking were inversely correlated with relative abundances of *Actinobacteria* and exhaled CO levels were inversely correlated with the abundances of *Cyanobacteria*. The number of cigarettes smoked per day was not associated with any bacterial taxa at the phylum level. Relative abundances of these bacteria were not associated with plasma TMAO, cardiovascular disease risk factors, or arterial measures. After a quit attempt, successful abstainers had significantly lower exhaled CO levels, but we did not observe significant changes in TMAO levels. We also did not observe between group differences in relative abundances of *Actinobacteria* or *Cyanobacteria*. Our primary observation was that relative abundances of *Bacteroidetes* increased and *Firmicutes* decreased in successful abstainers, however, we observed the reverse pattern in the continuing smoker group. We did not observe significant changes in alpha- or beta-diversity with smoking cessation. Overall, these findings represent relatively minor effects of smoking cessation on the microbiome.

An increased *Bacteroidetes:Firmicutes* ratio has been observed in several disease states that increase cardiovascular disease risk.[3,6,14-17] Increases in *Bacteroidetes* may increase lipopolysaccharide production which may upregulate inflammatory pathways.[18] *Bacteroidetes* also produce gut acetate, which is readily absorbed and can be used for cholesterol production.[14] Very few studies have examined the fecal microbiome of smokers after cessation. An observational study of 10 smokers suggested that smoking cessation led to a decrease in the *Bacteroidetes:Firmicutes* ratio.[6] A cross-sectional study that evaluated 758 men classified as never, current, or former smokers showed an increased *Bacteroidetes*: *Firmicutes* ratio in fecal microbiomes of current smokers compared to never smokers and former smokers, although there was no difference in this ratio between the never and former smokers groups.[20]

In a small study, smoking cessation was associated with a short-term increase in alpha-diversity.[15] We did not observe significant changes in alpha- or beta-diversity with smoking cessation.

This pilot study had several limitations including a relatively small sample size, though to our knowledge, this is the largest study of smoking and cessation on the fecal microbiota to date. Follow-up was 12 weeks and several subjects dropped out, as is common in studies of smoking cessation pharmacotherapy, so we can’t exclude a more long-term effect of smoking cessation. This study was limited to current smokers making a quit attempt, so we cannot compare our findings to non-smokers.

## CONCLUSIONS

Smoking cessation has minor effects on the composition of the gut microbiome. Among current smokers, relative abundances of the phylum *Actinobacteria* were associated inversely with pack-years of smoking and abundances of the phylum *Cyanobacteria* were associated directly with CO levels. In successful abstainers, relative abundances of the phyla *Bacteroidetes* increased and *Firmicutes* decreased, a pattern of uncertain clinical significance. We did not observe significant changes in alpha- or beta-diversity with smoking cessation. It is unclear if the proven health benefits of smoking cessation lead to salutary changes in the intestinal microbiota and if such changes affect cardiovascular disease risk.

## Data Availability

The data that support the findings of this study are available from the corresponding author upon reasonable request

---

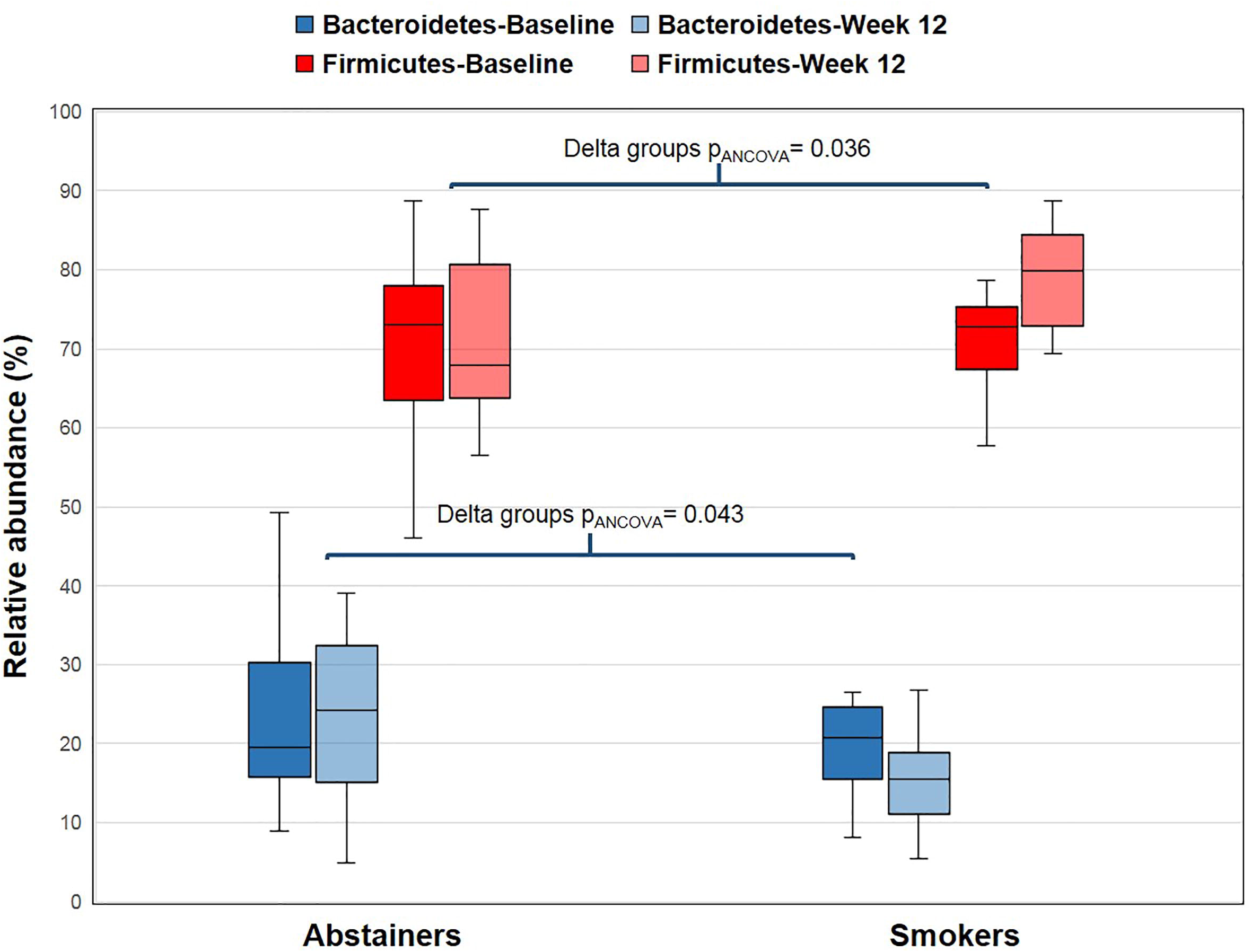

